# Alzheimer disease is (sometimes) highly heritable: Drivers of variation in heritability estimates for binary traits, a systematic review

**DOI:** 10.1101/2025.04.29.25326648

**Authors:** Shiying Liu, William S. Bush, Rufus Olusola Akinyemi, Goldie S. Byrd, Allison Mercedes Caban-Holt, Farid Rajabli, Christiane Reitz, Brian W. Kunkle, Giuseppe Tosto, Jeffery M. Vance, Margaret Pericak-Vance, Jonathan L. Haines, Scott M. Williams, Dana C. Crawford

**Affiliations:** Department of Population and Quantitative Health Sciences, Case Western Reserve University School of Medicine, Cleveland, OH, USA; Neuroscience and Ageing Research Unit, Institute for Advanced Medical Research and Training, College of Medicine, University of Ibadan, Ibadan, Oyo, Nigeria; Maya Angelou Center for Health Equity, Wake Forest School of Medicine, Winston-Salem, NC, USA; Department of Behavioral Science, College of Medicine and Sanders-Brown Center on Aging, University of Kentucky, Lexington, KY, USA; John P. Hussman Institute for Human Genomics, Dr. John T. Macdonald Foundation Department of Human Genetics, Department of Neurology, University of Miami Miller School of Medicine, Miami, FL, USA; Gertrude H. Sergievsky Center, Taub Institute for Research on Alzheimer’s Disease and the Aging Brain, Department of Neurology Columbia University, Department of Epidemiology, Columbia University, New York, NY, USA; The Taub Institute for Research on Alzheimer’s Disease and the Aging Brain, Columbia University Irving Medical Center, Columbia University; Department of Neurology, Columbia University Irving Medical Center, Columbia University; The Gertrude H. Sergievsky Center, College of Physicians and Surgeons, Columbia University Irving Medical Center, Columbia University, New York, NY USA; Dr. John T. Macdonald Foundation Department of Human Genetics, University of Miami Miller School of Medicine, Miami, Florida, USA

## Abstract

Estimating heritability has been fundamental in understanding the genetic contributions to complex disorders like late-onset Alzheimer’s disease (LOAD), and provides a rationale for identifying genetic factors associated with disease susceptibility. While numerous studies have established substantial genetic contribution for LOAD, the interpretation of heritability estimates remains challenging. These challenges are further complicated by the binary nature of LOAD status, where estimation and interpretation require additional considerations. Through a systematic review, we identified LOAD heritability estimates from 6 twin studies and 17 genome-wide association studies, all conducted in populations of European ancestry. We demonstrate that these heritability estimates for LOAD vary considerably. The variation reflects not only differences in study design and methodological approaches but also the underlying study population characteristics. Our findings indicate that commonly cited heritability estimates, often treated as universal values, should be interpreted within specific population contexts and methodological frameworks.

## Introduction

Heritability is commonly defined as the proportion of phenotypic variation attributable to genetics [1], a concept first described mathematically more than a century ago [2]. Early estimates of heritability for human traits were determined using family study design, while more contemporary estimates leverage population-scale studies with genome-wide genotyping or sequencing data [1,3,4]. Heritability estimates from the 1980s through the early 2000s were initially used to demonstrate that a trait of interest has a measurable genetic component [5], justifying the search for specific genes using either genetic linkage analyses or genetic association [6]. Almost 20 years into the era of genome-wide association studies (GWAS) and genome sequencing, estimates of heritability can now be used to estimate how much of a trait’s genetic component is yet to be discovered [7,8], thereby justifying additional genetic studies [9,10].

Both how heritability is estimated and how it is used have evolved over the past few decades. The literature on heritability study designs, statistical methods, and applications is deep, highlighting general strengths, weaknesses, and misinterpretations [3,4,11–13]. One major misinterpretation is that heritability is a fixed quantity [3,4]. In reality, heritability estimates are complex and dynamic, changing as a result of variable environmental effects and their interactions with genetic factors [14–17]. Study designs and statistical methodology also impact resulting heritability estimates. As an example, the heritability of the quantitative trait human height has been studied extensively for over a century. Classical family studies have consistently estimated the heritability of height at ∼ 80% [18], while the large-scale genetic studies of unrelated participants estimated that imputed genetic variation accounts for 56% of the observed phenotypic variance [19], illustrating the ceiling and floor of heritability estimates possible when different study designs and methodologies are deployed on different datasets from different contexts and samples.

Methods to estimate heritability were originally developed for quantitative traits. The concept was then extended and applied to binary traits such as late-onset Alzheimer’s disease (LOAD), a complex neurodegenerative disorder characterized by memory loss and dementia among adults ≥65 years of age [20,21]. One of the most highly cited estimates presents a very high heritability for LOAD (79%) [22], and while the identification of LOAD-associated genetic variants and genes has been successful [23–27], the range of LOAD heritability across a variety of contexts and methodologies and its implications for the discovery of new associating genes are not yet fully appreciated. It is worth highlighting that the methods for binary outcomes, such as LOAD, require additional steps for heritability estimation and cautious interpretation as compared to those for quantitative traits. This latter point we argue is not thoroughly described in the literature. In this systematic review, we examine LOAD heritability estimates, illustrate differences across study populations and study designs, and highlight differences based on assumptions and methodologies employed. Overall, we found that published LOAD heritability estimates indicate that generalizability is limited. In addition, the variation in reported LOAD heritability reveals population or cohort-specific variability that may be important in both identifying LOAD loci and designing interventions among populations.

## Methods

In this systematic review, we focus on narrow sense (h^2^) heritability of LOAD as this is generally the measure presented in the literature. Heritability can also be estimated as broad-sense (H^2^) heritability. H^2^ captures the total genetic variance, which includes additive (A) and dominance (D) effects, as well as the epistatic effect (I). In contrast, h^2^ only defines the additive genetic component. While closely related, these two metrics describe distinct but overlapping aspects of genetic influence on phenotypic variation within a population. Distinguishing these forms is important, as each has different implications and requires flexible interpretation in research contexts [11,28].

### Data collection

Data were collected from the primary literature and included original, peer-reviewed studies of LOAD narrow sense heritability. Our primary literature search was conducted using PubMed, the National Library of Medicine’s free resource for the search and retrieval of citations and abstracts from MEDLINE and PubMed Central (PMC) databases [29]. We formulated the search query as “Alzheimer’s disease” AND “heritability,” tailored to PubMed’s search functions, to identify articles that mentioned heritability values for LOAD. This initial search, conducted in April 2024, yielded 624 articles, including both research and review papers.

In addition to the initial search, we explored other resources to ensure that our review was comprehensive and not limited by PubMed or the corresponding search terms we used. These included relevant publications identified from citations of published twin registries focusing on LOAD, along with citations within and publications citing the recent large LOAD GWAS captured in the initial search results. This “backward searching” added 52 articles, and these articles were combined with our initial search results and screened using the approaches as described below.

To finalize the pool of articles to include in this systematic review, we applied a multi-step filtering process to identify relevant studies from the initial search, excluding studies that only mention or restate published heritability values. Manual screening of the titles and abstracts of all 676 papers was first performed to exclude articles that were irrelevant, such as those not focusing on LOAD. This initial screening eliminated 64 papers, leaving 612 papers to consider. Of these 612, 23 were excluded because they could not be retrieved through institutional access or were non-English publications. For the remaining 589 articles, we conducted a comprehensive screening of both abstracts and full texts to identify studies that involved original research on the heritability of LOAD. We first removed studies that only referenced the heritability value(s) of LOAD without calculating their own estimates (n=512; Fig 1). We then performed an in-depth full-text examination of the remaining 77 papers to determine if they met our criteria for conducting original research on heritability estimation for LOAD. We defined original research as studies with detailed methods for computing heritability, including details of the study design, modeling process, and study population. Manuscripts without sufficient detail were excluded from further consideration. For example, one retrieved twin study lacked explicit details on how the LOAD heritability estimates were derived [30]. A parent-offspring study was also excluded due to inaccessible supplementary materials that described both the study population and the detailed methodology [31]. In total, 54 manuscripts were excluded at this step (Fig 1), leaving 23 peer-reviewed manuscripts with sufficient detail for this review.

**Fig 1.**
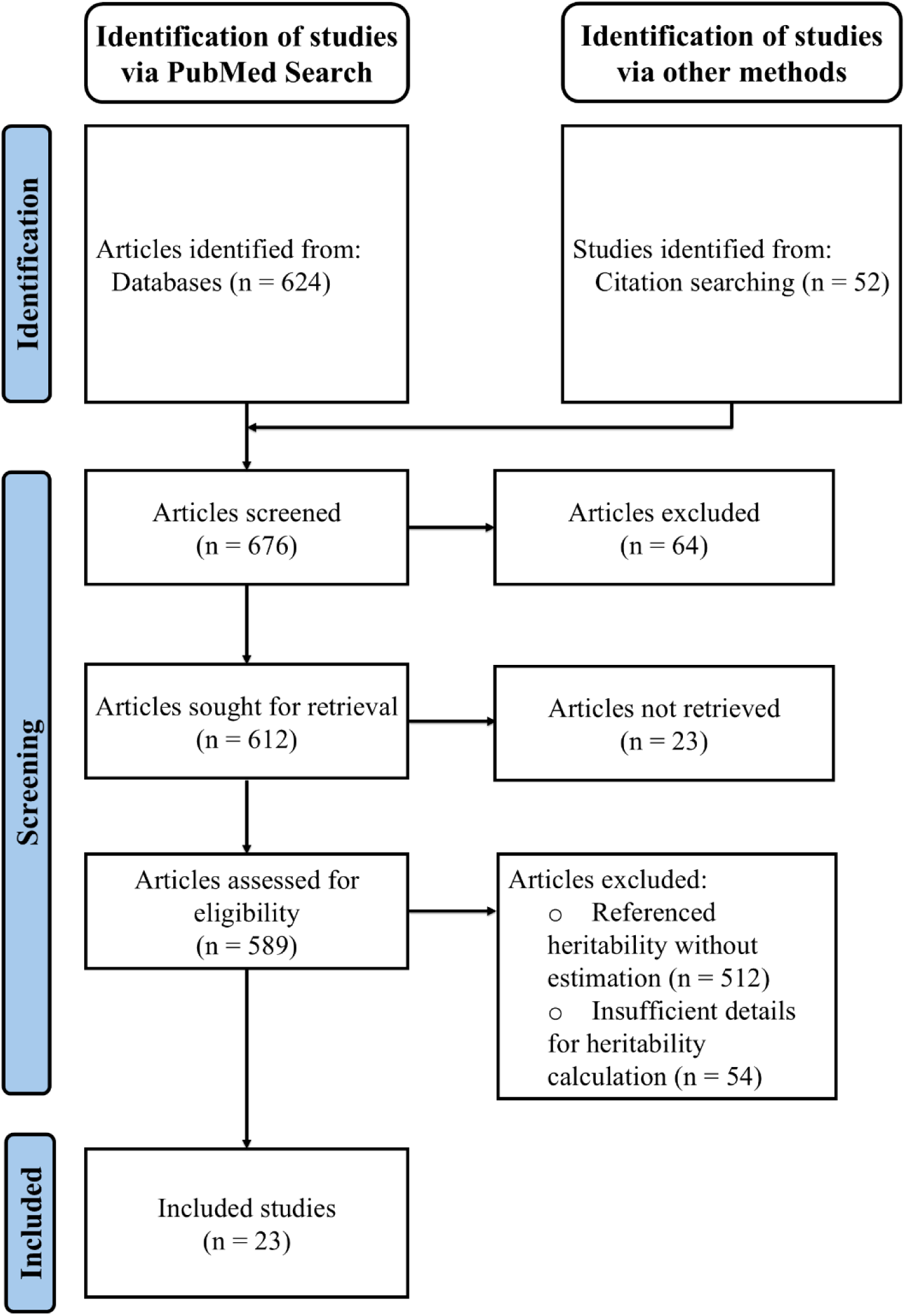
PRISMA flow diagram. Created following the PRIMSMA 2020 guidelines [32].

### Characteristics of the Literature

We categorized the 23 eligible studies based on the study design and methods employed. The evaluation included the data sources, the study design employed in the original studies, the assumptions embedded in each approach, and the populations to which these estimates are intended to be generalized [8,19,28,33–36]. Heritability estimation methods can be categorized into two primary groups: 1) family-based methods, rooted in studies of related individuals, such as twins and extended pedigrees, without necessarily requiring genomic data, and 2) single nucleotide polymorphism (SNP)-based methods, consisting of population-based methods leveraging genome-wide data from large samples of often unrelated individuals. Based on this, we examined the articles, focusing on their methodologies, and stratified them into twin-based and SNP-based heritability estimates. Among the 23 articles passing the criteria, seven were based on twin studies, while 16 were SNP based studies.

This review has not been registered, and no protocol was prepared.

## Results

We identified a total of 23 relevant studies published between 1997 and 2023, including both twin-based and SNP-based studies. In addition to the heritability estimate and standard error, for each study the following essential information was collected for all studies: study population characteristics, sample size, case criteria, and specific estimation methods (Tables 1-3). For SNP-based studies, we also collected number of SNPs, included covariates, summary statistics dataset reference, and prevalence used for liability transformation (Tables 2 and 3). The heritability estimates for LOAD across different study designs and populations ranged from 3.1% [26] to 79% [22]. These studies primarily focused on participants of European ancestry, with a mean age mostly greater than 70 years, except for one study leveraging the National Research Council Registry of Aging Twin Veterans that has an average age of 63.1 years [37]. The sample sizes varied considerably, ranging from 38 twin pairs [38] to 761,704 participants in a SNP-based study [26]. Compared with SNP-based estimates, heritability estimates from family-based studies generally provided higher values, representing the likely ceiling of trait heritability [8]. SNP-based estimates correspond to the proportion of genetic variance explained by SNPs assayed, imputed, or tagged and therefore are unlikely to capture the totality of genetic effects. Given the fundamental differences of what is being measured, we present our results in two main categories: twin-based heritability estimates and SNP-based heritability estimates. We then address details within each category by examining specific methodologies and their impact on the resulting heritability values, providing a comprehensive view of LOAD heritability across diverse study designs and populations.

**Table 1.**
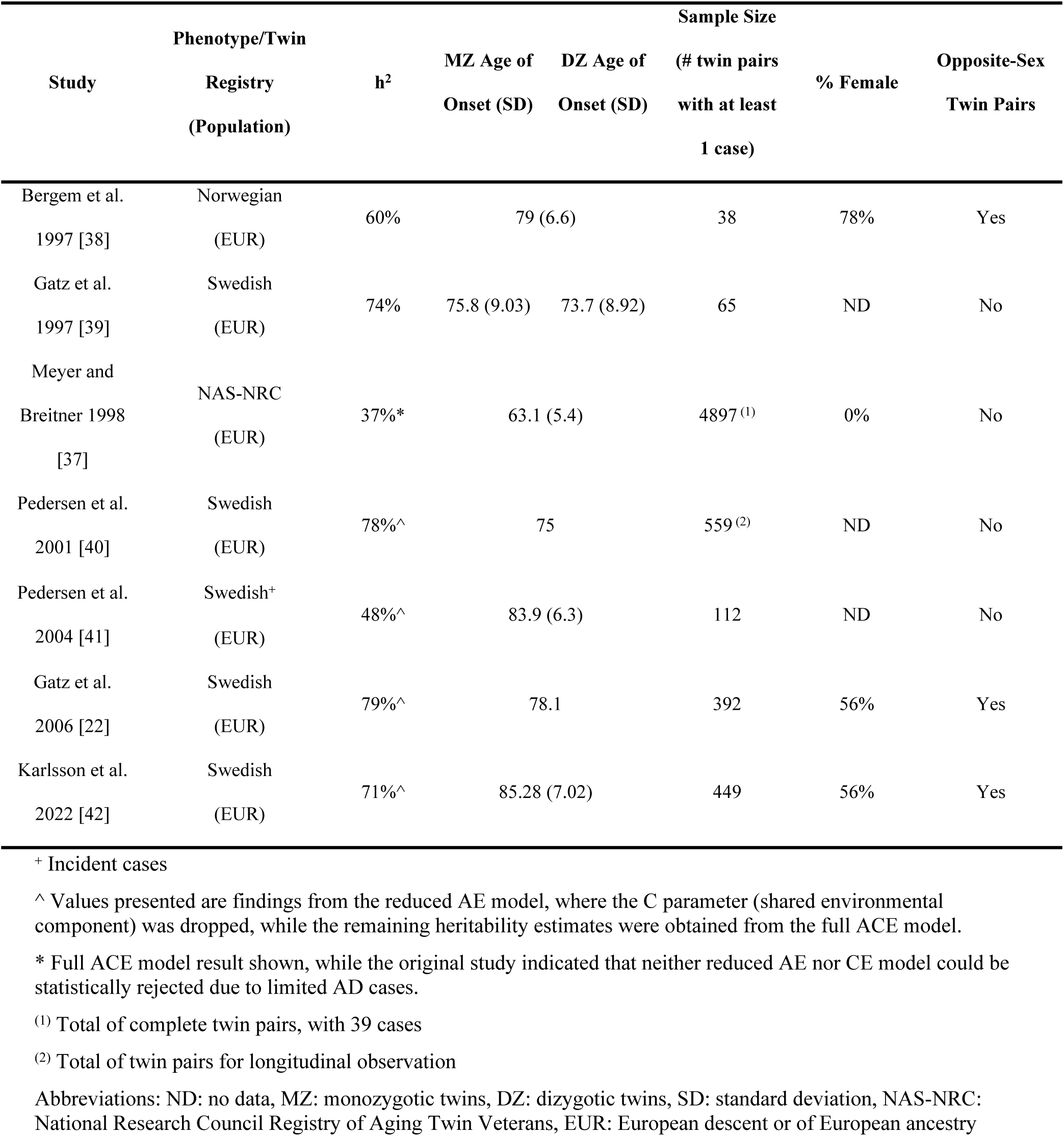
Twin Study-based LOAD heritability studies.

| Study | Phenotype/Twin Registry<br>(Population) | h<sup>2</sup> | MZ Age of Onset (SD) | DZ Age of Onset (SD) | Sample Size |  | Opposite-Sex Twin Pairs |
| --- | --- | --- | --- | --- | --- | --- | --- |
|  |  |  |  |  | (# twin pairs with at least 1 case) | % Female |  |
| Bergem et al. 1997 [38] | Norwegian (EUR) | 60% | 79 (6.6) |  | 38 | 78% | Yes |
| Gatz et al. 1997 [39] | Swedish (EUR) | 74% | 75.8 (9.03) | 73.7 (8.92) | 65 | ND | No |
| Meyer and Breitner 1998 [37] | NAS-NRC (EUR) | 37%* | 63.1 (5.4) |  | 4897<sup>(1)</sup> | 0% | No |
| Pedersen et al. 2001 [40] | Swedish (EUR) | 78%^ | 75 |  | 559<sup>(2)</sup> | ND | No |
| Pedersen et al. 2004 [41] | Swedish+ (EUR) | 48%^ | 83.9 (6.3) |  | 112 | ND | No |
| Gatz et al. 2006 [22] | Swedish (EUR) | 79%^ | 78.1 |  | 392 | 56% | Yes |
| Karlsson et al. 2022 [42] | Swedish (EUR) | 71%^ | 85.28 (7.02) |  | 449 | 56% | Yes |

+ Incident cases
^ Values presented are findings from the reduced AE model, where the C parameter (shared environmental component) was dropped, while the remaining heritability estimates were obtained from the full ACE model.
\* Full ACE model result shown, while the original study indicated that neither reduced AE nor CE model could be statistically rejected due to limited AD cases.
<sup>(1)</sup> Total of complete twin pairs, with 39 cases
<sup>(2)</sup> Total of twin pairs for longitudinal observation
Abbreviations: ND: no data, MZ: monozygotic twins, DZ: dizygotic twins, SD: standard deviation, NAS-NRC: National Research Council Registry of Aging Twin Veterans, EUR: European descent or of European ancestry

**Table 2.**
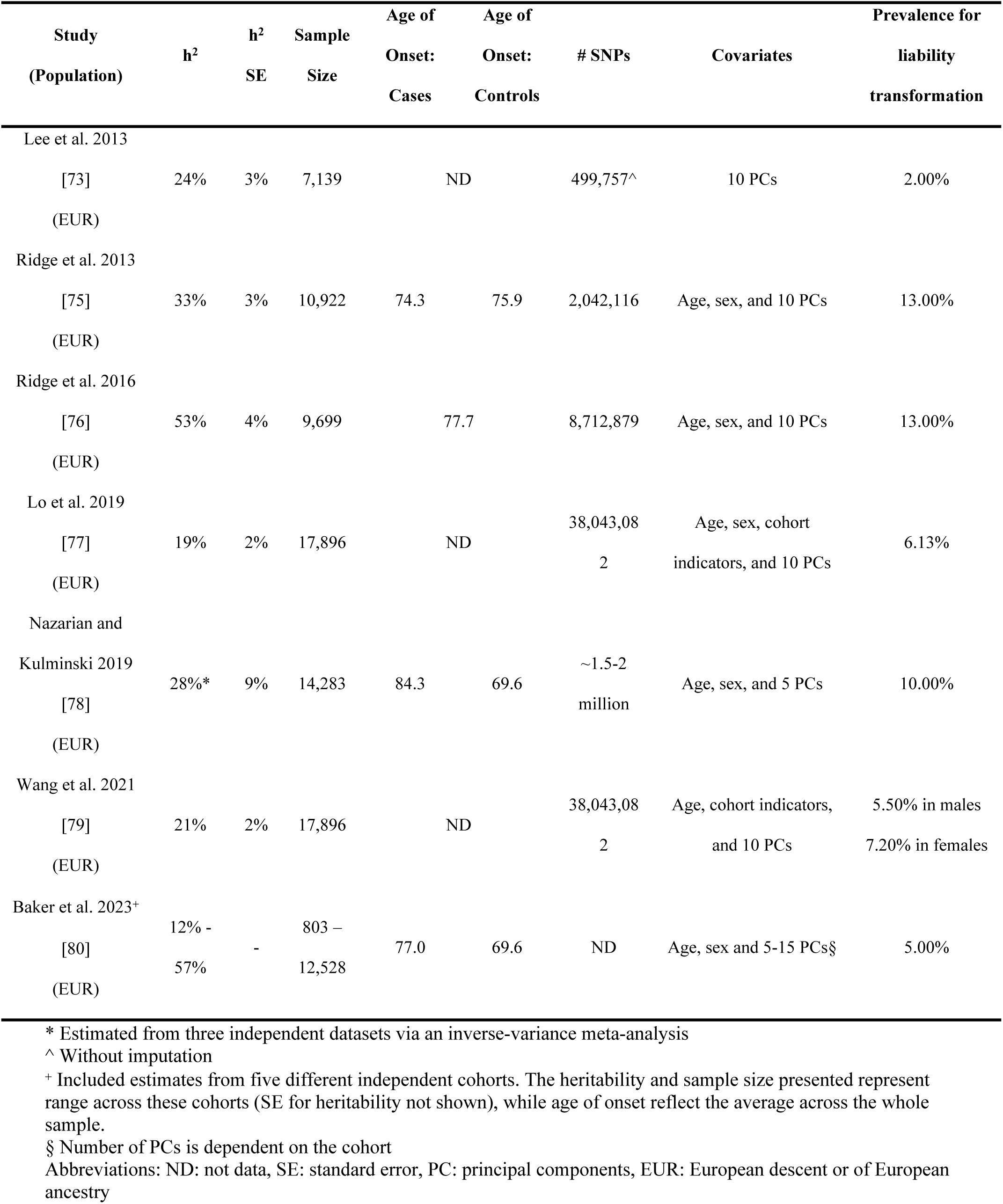
SNP-based LOAD heritability studies using GCTA-GREML.

### Twin-based Heritability

Seven studies using the family-based approach met our inclusion criteria, all of which were based on twin studies. Despite similar study designs and similar estimation approaches, LOAD heritability estimates based on twin studies vary widely, ranging from 37% to 79% (Table 1; Fig 2). We further investigated the potential reasons that could lead to the differences among the estimated values, including variations in twin registries, study population characteristics, and LOAD phenotyping.

**Fig 2.**
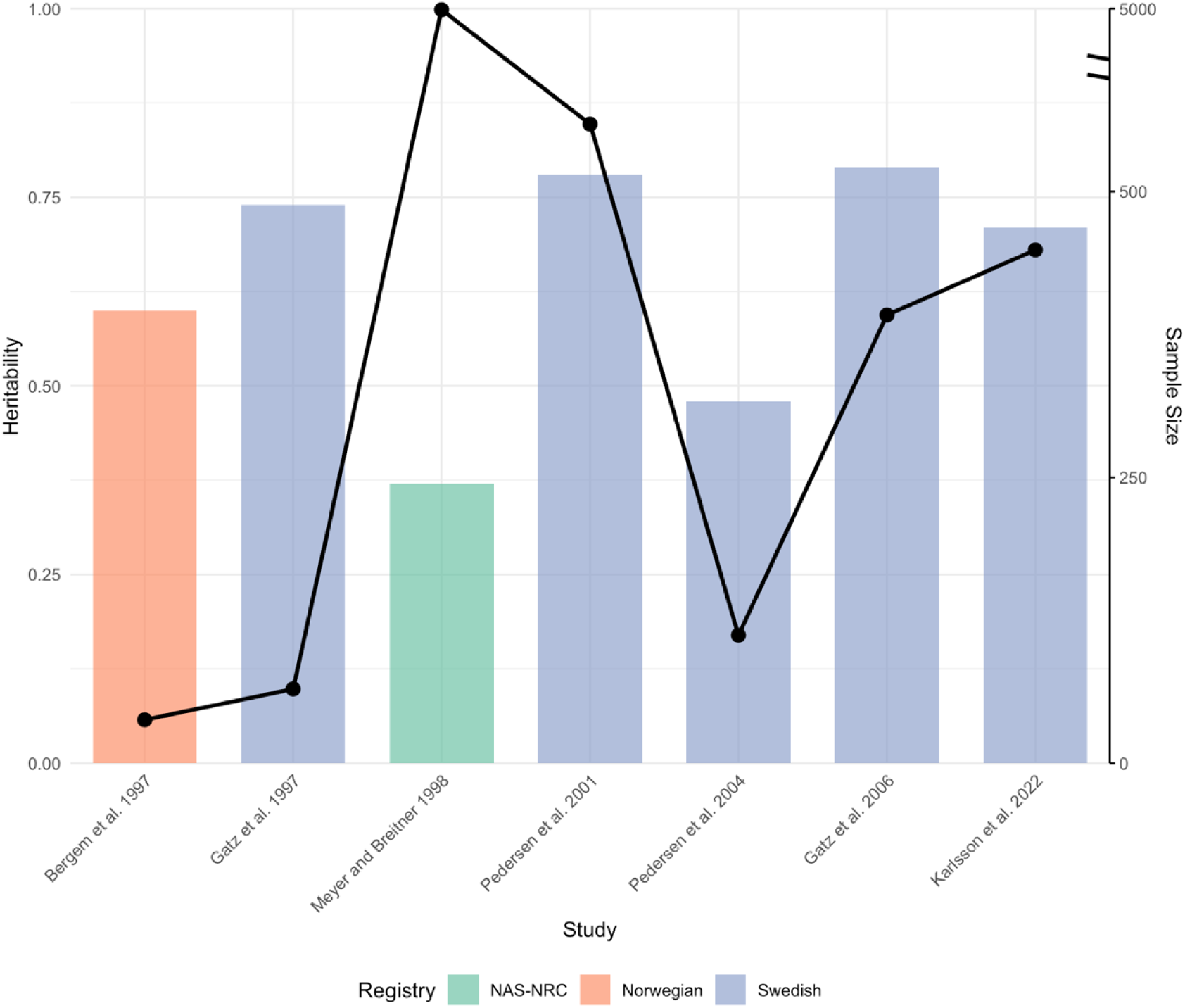
Twin study-based heritability estimates and sample sizes across studies. The figure presents the heritability estimates (bars, left y-axis) and corresponding sample sizes (line, right y-axis) for studies arranged chronologically along the x-axis. Each bar represents a single study, with colors denoting different twin registries utilized. There is a break mark, indicated by a double slash, on the right y-axis between 500 and 4850 to accommodate the wide range of values for sample sizes.

### Twin Registries

The majority of the identified studies used the Swedish Twin Registry (STR) [43], with two exceptions: one study leveraged the Norwegian Twin Registry (NTR) [44] and another the National Research Council Registry of Aging Twin Veterans (NAS-NRC), also from Norway [45]. The NTR study had the smallest sample size (n = 38 twin pairs with AD diagnosis) [38], a size expected given the study formulated the target probands via cross-referencing the cognitively impaired elderly born between 1895-1925 within the twin registry. The NAS-NRC panel, established in the mid-1950s, focused on white male twins [46]. The registry’s focus on white males and the high non-participation rate of (24%) of the LOAD study [37] potentially makes its heritability estimates less representative compared with other estimates [45].

The STR-based LOAD heritability studies were more complex than those from the two registries discussed above, involving longitudinal efforts and variability within sub-studies. Three studies leveraged samples from the Study of Dementia in Swedish Twins [39] based on the Swedish Adoption/Twin Study of Aging (SATSA), a subset of the population-based STR with detailed enrollment criteria [47]. These studies included twins drawn from STR with variations in the inclusion criteria across studies, resulting in varying numbers of twin pairs in the analysis. In addition to the SATSA panel, Pedersen et al (2004) also leveraged the Origins of Variance in the Oldest Old: Octogenarian Twins (OCTO-Twin) [48] that incorporated longitudinal observations into the study, resulting in 662 twin pairs without symptoms with follow-ups spanning on average five years [41]. The sample size was further enriched with the introduction of the Study of Dementia in Swedish Twins (HARMONY) initiated in the year 1998 [49]. Prior to the HARMONY study, STR assessments were not comprehensive for dementia, limiting sample sizes. A larger sample size was achieved in the Gatz et al (2006) study (n = 392 pairs with at least 1 AD case) that employed a two-phase phenotype scheme where all participants were initially screened for cognitive dysfunction and suspected AD cases were followed up with complete clinical diagnostic evaluations. This STR LOAD heritability study also included opposite-sex twin pairs [22]. More recent efforts have further expanded the use of STR data by incorporating four STR sub-studies (SATSA, OCTO-Twin, HARMONY, and Aging in Women and Men) [42]. The inclusion of Aging in Women and Men (also known as GENDER) added value by introducing opposite-sex twin pairs to LOAD heritability research [50].

Other twin registries exist that are not included in this review. For example, the Finnish Twin Registry study by Räihä et al. (1996) [51] was also mentioned by Pederson et al (2001) as part of their exploration of LOAD heritability using a single-threshold model, although it was not the primary focus of their research [40]. The original study included same-sex twin pairs from Finland born before the year 1958, with disease status identified through the linkage to the Hospital Discharge Register, leading to a total of 94 AD affected out of 178 twin individuals [51]. Incomplete record linkage could potentially explain the lower heritability estimate of 63% reported by Pedersen et al. (2001) for the Finnish Twin Registry study [40].

### Twin Registry Characteristics

The study populations differed in numerous characteristics that may influence measures of heritability. Notable variation in participants’ ages, a demographic strongly correlated with LOAD prevalence [52], were observed across studies. The twin veterans study had an average age of 63.1 years (SD: 5.4) [37]. This younger sample might not have fully manifested LOAD symptoms at the time of assessment, potentially contributing to a relatively lower heritability estimate (h^2^ = 37%). In contrast, Pedersen et al. (2004) observed a much later average age of 83.9 years (SD: 6.3) [41] and higher heritability (48%; Table 1) that could be attributed to their longitudinal study design, where twin pairs aged 52 to 98 were followed for approximately 5 years. The Karlsson study in 2022, leveraging enriched samples from the STR, benefitted from a longer follow-up (through 2016) resulting in an average age of 85.28 years (SD: 7.0) and higher heritability (71%) compared with both Meyer and Breitner (1988) and Pederson et al (2004) [42]. These age variations highlight the importance of considering age, and when possible, birth cohort, of the participants in LOAD studies to capture a more comprehensive picture of its heritability.

Several studies also explicitly indicated the inclusion of different sex twin pairs. LOAD disproportionally affects females, with higher lifetime risk and sex-specific risk factors observed in previous studies [53,54]. Moreover, prior research indicates that genetic components may manifest differently in males and females [55]. Bergem et al. (1997) was the first study we identified that included twins of differing sex; however, it was limited by a small proportion of male-female twins (33% among the dizygotic twin pairs). Later studies leveraging HARMONY, as described above, achieved a more balanced composition of like- and unlike-sex twin pairs; the former study did not identify differences in heritability across sexes, while the latter also incorporated additional opposite-sex twin pairs drawn from GENDER, representing 21% of the total sample.

One study stands out for its focus on incident LOAD cases rather than prevalent cases [41]. While most studies examine prevalent cases to estimate heritability, this study incorporating two longitudinal twin studies investigated the relative importance of genetic and environmental impact in disease development across different age of onset groups. The unique study design yielded an overall heritability estimate of 48%, with age-stratified estimates of 59% for onset before age 80 and 40% for onset after age 80. These estimates are generally lower than those obtained from studies of prevalent cases (Table 1), revealing the potential impact of study design on heritability estimates.

### Twin Study Modeling Strategies

A straightforward method for heritability estimation is Falconer’s formula [*h*^2^ = 2(*r*_MZ_ ― *r*_DZ_)], which relies on the phenotypic correlation between twin pairs [56]. Bergem et al. (1997) applied Falconer’s formula to estimate the heritability of AD, incorporating a series of probability percentages for positive cases, and obtained heritability estimates ranging from 55% to 61% [38]. An advantage of phenotypic correlation is its ease in interpretability. However, this approach relies heavily on the assumption of equal shared environmental variance and lacks the ability to address more complex aspects of genetic analysis, such as model performance assessment, incorporation of extended family data, and investigation of gene-by-environment interactions, potentially leading to oversimplified estimates that do not fully capture the genetic architecture of complex traits like LOAD.

With advances in analytical techniques and software for effective data handling, more sophisticated modeling approaches have been developed to estimate variance components using information from twin studies. Among these methods, structural equation modeling (SEM) has become frequently employed, with model fitting relying on maximum likelihood estimation.

SEM offers greater flexibility in determining the contributions of additive genetic (A), shared environmental (C), and unique environmental (E) components, collectively forming the ACE model, while also allowing for the incorporation of covariates into the analysis, providing a more comprehensive understanding of the factors influencing heritability [11,57]. Typically, these studies involve testing both the full ACE model and reduced models, successively dropping either A or C, to identify the best-fitting model along with the corresponding parameter estimations [22,37,39–42]. Model performance is generally assessed using indicators such as the Akaike Information Criterion (AIC) and chi-square test of difference, while some studies also incorporate critical information, such as consistency with observed correlations and concordance estimates, into the determination of the best-fitting model [39,41].

The majority of the heritability estimates presented in LOAD twin studies (Table 1) originated from the AE model, which excludes shared environmental variance, assuming that additive genetic components and unique environmental impact are the primary contributors to LOAD risk. Exceptions to this approach include Gatz et al. (1997) who reported that the full ACE model performed best, estimating heritability at 74% and attributing 24% to the shared environmental effects [39]. Meyer et al. (1998) found neither the AE nor CE models could be rejected compared to the full model, yielding heritability estimates of 74% in the AE model and 37% in the full model [37]. Collectively, these studies demonstrated the importance of considering multiple models and interpreting results cautiously. The preference for the AE model in the LOAD twin studies does not necessarily negate the relevance of shared environmental factors. Instead, it indicates their contribution may be minor or more difficult to detect given current methodologies, emphasizing the need for larger studies and sophisticated modeling approaches to fully elucidate the complex interplay of factors contributing to disease risks.

In heritability studies of LOAD, analyzing binary traits (affected vs. unaffected) presents unique challenges compared to quantitative traits [58,59]. Dichotomous outcomes necessitate a specialized approach to estimating variance component partitions, typically via the assumption of a latent, normally distributed liability for the trait. Disease status is then determined by a threshold, often corresponding to the population disease prevalence, with individuals above this threshold considered affected, while those below as unaffected. This liability threshold model is necessary for incorporating binary outcomes into a framework suitable for heritability analysis. Thus, it is important to use the appropriate threshold(s) in the study, not only to address the ascertainment bias embedded in the twin study design but also to accurately reflect the population characteristics for age-dependent diseases, like LOAD.

The complexity of this process is evident in the approaches adopted in different studies. Some studies leveraged previously published epidemiological data to inform the liability threshold. Bergem et al. (1997) utilized the published population prevalence of LOAD, setting the prevalence at 10% in their analysis based on previous studies with similar age distributions [60,61]. In contrast, several other studies [22,37,39–41] relied on the estimated prevalence or incidence rates derived from their study samples. While this method potentially offers specificity to the study population, it makes it difficult to compare heritability estimates across studies.

Given the challenges of using prevalence or incidence, some studies focused on a threshold modeling process for LOAD to better capture the potential right-censoring in the data, along with the accurate representation of study samples. This methodology moves beyond the single threshold model used in earlier studies [38,39] to more sophisticated approaches that address the relationship between age of onset and disease liability, such as the implementation of multiple threshold models. Meyer et al. (1998) and Pedersen et al. (2001) incorporated the five-year age groups for monozygotic and dizygotic twin groups, assigning multiple thresholds corresponding to the specific prevalences estimated in these stratified samples [37,40]. Pedersen et al. (2001) further refined this approach by incorporating an additional age group or bin to distinguish censored observations and by testing models with both population-based and sample-specific prevalence estimates [40]. Pedersen et al. (2004) also incorporated differing incidence rates within two age groups (below and above 80 years old) in their heritability analysis. While this approach has the potential to offer a more nuanced understanding of age-related genetic influences on disease onset, no differences were observed in estimates for twins <80 years of age compared with twins ≥80 years of age. [41].

One study not only employed biometrical analysis as discussed above but also incorporated a sex-limitation twin model, leveraging data from a reasonable number of different-sex twin pairs [22]. Unlike previous studies that had a single major stratum based on twin similarity (e.g., monozygotic versus dizygotic), Gatz et al. allowed for estimated thresholds to differ both by sex and age of twins. This more complex modeling offered more insights into variance partitioning, enabling the estimation of sex-specific effects and demonstrating the lack of differences in LOAD heritability between men and women.

### Twin Study Meta-Analyses

In addition to the seven twin-based heritability studies detailed here, two meta-analyses based on family data from individual studies exist in the literature. In one meta-analysis, a weighted mean heritability was estimated based on five previously published twin studies [62]. Among the studies meeting their inclusion criteria, four [22,37–39] are detailed in Table 1 whereas the additional included twin study is from the Finnish twin registry [51]. By weighing the number of twin pairs where at least one is affected by the disorder, the analysis yielded a heritability estimate of 75%. Another meta-analysis of twin correlations from seven studies is available in the literature [63] that includes all of the twin studies described in Table 1 except Karlsson et al. 2022, which was published later. Overall, monozygotic twins for dementia in Alzheimer’s disease had a higher correlation (0.86) compared with dizygotic twins (0.50) (MaTCH) [63]. Using ACE models, the meta-analysis yielded a heritability of 63% for all twins and 59% for the same sex twins [63]. Applying a different method, least squares models based on Falconer’s formula, to the same data led to a higher heritability estimate of 71% for the same-sex twins [63].

### SNP-Based Heritability

Compared with study designs requiring related individuals, population-based approaches for heritability estimation have gained popularity in recent years largely due to the widespread availability of genome-wide SNP data from large case-control studies as well as the advancements in statistical methodologies [36,64–67]. Unlike family-based studies, the population-based approaches utilize the realized genetic matrices computed from the genetic data of large cohorts, circumventing the need for specific family designs and thus broadening the scope for heritability estimation [68]. SNP-based heritability refers to the estimated heritability that is attributable to the assayed or imputed SNPs associated with a complex trait or outcome. Multiple methods have been developed to estimate SNP-based heritability. Some require individual-level data while others require only summary statistics [69].

### GCTA

Genome-wide complex trait analysis (GCTA) is one of the most commonly used methods that leverages individual-level data while relying on different algorithms [70,71]. GCTA employs linear mixed models (LMM) with a normality assumption for the residuals and partitions the phenotypic variance into variance components, leveraging the genetic relatedness matrix (GRM) constructed from the genetic data. The heritability can then be estimated through the genome-based restricted maximum likelihood (GREML) procedure. We identified seven LOAD heritability studies that employed this method (Table 2).

The first LOAD GCTA heritability estimates were published in 2013 (Table 2; Fig 3). The resulting LOAD heritability estimate of 24% was based on a sample of 7,139 participants (3,290 cases, 3,849 controls) that included both elderly screened and population controls [72,73]. Concurrently, Ridge et al. (2013) leveraged the Alzheimer’s Disease Genetics Consortium (ADGC) dataset [74], including 10,922 individuals (5,708 cases, 5,214 controls), and estimated a higher heritability estimate of 33% [75]. The ADGC dataset has since continued to expand, facilitating more comprehensive analyses of LOAD. A 2016 update included data from 30 studies within the ADGC, and the heritability estimate increased to 53%. This study included only 9,699 individuals (3,877 cases, 5,822 controls) [76]. This sample size was smaller in this latter study due to the requirement for non-missing data across 21 known AD genes, resulting in a higher LOAD heritability estimate compared with the 2013 analysis and highlighting the impact of the quality control (QC) process and demand for complete data at known LOAD risk loci. Inclusion of more known LOAD genes increased the heritability, indicating that these genes have substantial impact on overall estimates.

**Fig 3.**
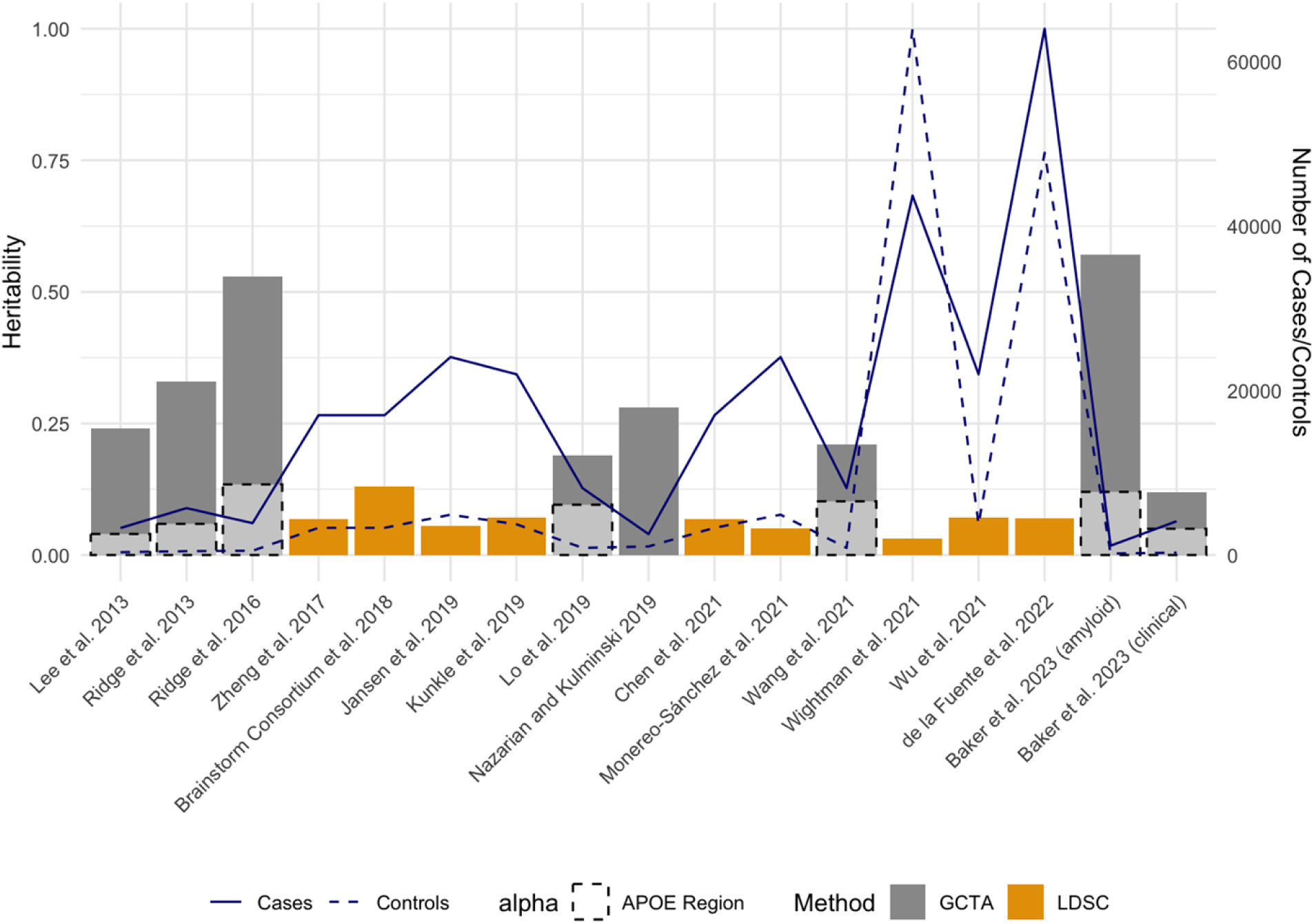
SNP-based heritability and sample sizes across studies. The figure illustrates SNP-based heritability estimates and sample sizes for different studies, ordered by year of publication and author. Bars represent heritability estimates obtained from each study, with different colors indicating the methods used to estimate heritability (GCTA or LDSC). The solid line demonstrates the number of cases in each study, while the dashed line represents the number of controls.

Later and larger two-phase ADGC datasets focusing on age- and sex-specific heritability utilized samples of up to 17,896 participants [77,79]. Unlike the smaller twin study of Pederson et al (2004) that found no difference in LOAD heritability by age, Lo et al (2019) with a much larger sample size of unrelated individuals estimated a higher heritability for participants >80 years of age (n=5,198; h^2^=24.1%) compared with those aged 60-69 years of age (n=12,698; h^2^=16.9%). Differences by sex were subtler with a slightly higher LOAD heritability among women (21.5%) compared with men (19.5%) [79]. Overall, variation in study design, case definition, and sample characteristics can greatly impact heritability estimates, necessitating a more cautious interpretation of these findings.

The evolution of LOAD genetic studies has been marked not only by increasing sample sizes but also by a substantial enhancement in marker density. This improvement can be attributed to advancements in both genotyping technologies and imputation techniques, significantly impacting the comprehensiveness of SNP-based heritability estimates. The relatively lower heritability estimation (h^2^ = 24%) obtained by Lee et al (2013) can be partially attributed to the less dense genotype data, utilizing only 499,757 SNPs [73]. This limitation in marker density likely constrained its ability to fully capture SNP-based heritability, which was also supported by the observed decrease in heritability estimates when more stringent QC processes were applied. As opposed to being solely dependent on the directly genotyped data, subsequent studies have leveraged the power of imputation, hugely increasing the number of genetic variants analyzed. In the same year, Ridge et al. (2013) exemplified this leap forward by employing data imputed against the HapMap phase II (release 22) reference panel [81], resulting in 2,042,116 SNPs after quality control, a substantial increase compared to non-imputed datasets that all had fewer than 500,000 SNPs [75]. The trend towards higher marker density has been further accelerated by advances in imputation reference panels. The adoption of the 1000 Genomes Project reference panel [82], offering more comprehensive coverage, has enabled even denser imputation. Ridge et al. (2016) utilized 8,712,879 SNPs in their analysis, while for more recent studies leveraging the two-phase ADGC data, up to 38 million SNPs were incorporated in the analysis [77,79]. The substantial increase in marker density, from ∼500,000 to 38 million SNPs, represents a significant methodological advancement in LOAD genetic studies albeit with new analytical challenges, including more stringent requirements for QC processes and increased computational demands. While the differences in marker density could potentially explain some of the variations in heritability estimates across studies, they alone do not explain the observation that the less dense dataset of Ridge et al (2016) has a much higher heritability (53%) compared with either Lo et al (19%; 2019) or Wang et al (21%; 2021).

Analyses with fewer samples and SNPs provided higher estimates, but the analyses differed in ways that may inform our understanding of heritability estimates. For example, inclusion of specific covariates and variable assumed prevalences, among other variables such as marker density (**Error! Reference source not found.**), complicate comparisons across studies and heritability expectations. As noted above, the GCTA approach that uses LMM allows for the inclusion of covariates as fixed effects, enabling more accurate heritability estimates for LOAD. Age and sex, well-established factors affecting LOAD risk, have been consistently included as covariates in most studies, with the notable exception of Lee et al. (2013) [73]. Principal components (PCs) are universally recognized as crucial for adjusting population structure in genetic studies [83]; however, there is considerable variation in both the number of PCs included in the analyses and how the PCs themselves are calculated. While most studies incorporate 10 PCs [73,75–77,79], Nazarian and Kulminski (2019) used only the first five PCs (Nazarian and Kulminski 2019), and Baker et al (2023) employed a flexible approach where the number of PCs was cohort specific [80]. PCs can be calculated within individual cohorts, as demonstrated by Lo et al. (2019), or using the entire combined dataset as did Ridge et al [76]. These methodological heterogeneities may also contribute to differences observed across heritability estimates.

Methodological heterogeneity when using large collaborative studies to estimate SNP-based heritability with GCTA is not limited to PCs. In practice, large collaborative studies are made up of different cohorts that have undergone varied ascertainment processes and genotyping strategies. While increased sample size and diversity are valuable, they come at the cost of potential heterogeneity across cohorts. Lo et al. (2019) made a significant methodological advance by incorporating cohort indicators into their heritability estimation [77]. This approach yielded a lower estimate of 19% for LOAD compared to previous studies. Their sensitivity analysis showed an increase to 32% when cohort indicators were removed, revealing a substantial cohort effect. Wang et al. (2021) similarly incorporated cohort indicators in their analysis of 2-phase ADGC data, obtaining an expected comparable heritability estimate of 21% [79]. While GCTA-based approaches have significantly advanced our understanding of LOAD heritability, they underscore the need for cautious interpretation of estimates due to methodological variation, particularly in covariate handling, highlighting the importance of continued refinement of analytical strategies. We also cannot ignore the possibility that cohorts within a single study actually have variation in true heritability due to unmeasured environmental parameters.

LOAD phenotyping approaches across studies used to estimate heritability are highly heterogeneous. The majority of heritability estimates, particularly those utilizing data from ADGC, rely on a combination of clinical diagnosis, histopathologic findings, and in some cases, biomarkers. Clinical diagnosis have inherent misdiagnosis rates [84] that can affect heritability estimates. Baker et al (2023) illustrated this by demonstrating that the histopathologically confirmed LOAD yielded higher heritability estimates (31% - 57%) compared to clinically diagnosed LOAD (12% - 32%) when applying a consistent model with a 5% liability threshold to five independent cohorts [80]. Notably, within the Amsterdam Dementia Cohort, using the amyloid-confirmed cases showed a heritability estimate of 57%, while the clinical diagnosed cases from the same population yielded an estimate of 25%. This marked difference emphasizes the significant impact of diagnostic criteria on heritability estimation, potentially helping explain some of the observed variability in heritability across studies.

SNP-based heritability estimates of LOAD must address *APOE*, the major genetic risk factor for LOAD [23]. Compared with other complex trait-associated genetic variation, *APOE* has an outsized effect and alone contributes substantially to LOAD heritability, accounting for an estimated 4% to 13.42% of the total phenotypic variance (Fig 3) [73,75–77,79]. The lowest heritability estimate (4%) was obtained using proxy SNPs for *APOE* [73], and although not significantly different from the proxy-based estimate, two higher heritability estimates for *APOE* were obtained when the *APOE* ε2 and ε4 alleles were directly genotyped [75,76]. In contrast to the SNP partitioning method, some studies employed an approach leveraging the best linear unbiased prediction (BLUP) and involved regressing on the number of *APOE* ε4 alleles to estimate heritability attributable specifically to the *APOE* ε4 alleles [77,79]. Regardless of the variability, published estimates confirm that *APOE* explains a substantial portion of the genetic risk for LOAD.

### LDSC

With the proliferation of large-scale GWAS, and their accompanying summary statistics, methods to estimate heritability such as linkage disequilibrium (LD) score regression (LDSC) have become popular. LDSC only requires summary-level data and involves the process of LD score calculations using the appropriate reference panel for each variant. These calculations can then be incorporated into a regression model where the observed GWAS summary statistics are regressed against the pre-computed LD scores to compute the heritability estimation [85,86]. While convenient and in some cases the only option for difficult-to-access datasets, such methods typically lead to an underestimation of SNP-based heritability compared with those derived from individual-level data [36,87] as evident for LOAD (Tables 2 and 3). We identified nine LOAD heritability studies that employed LDSC (Table 3).

**Table 3.**
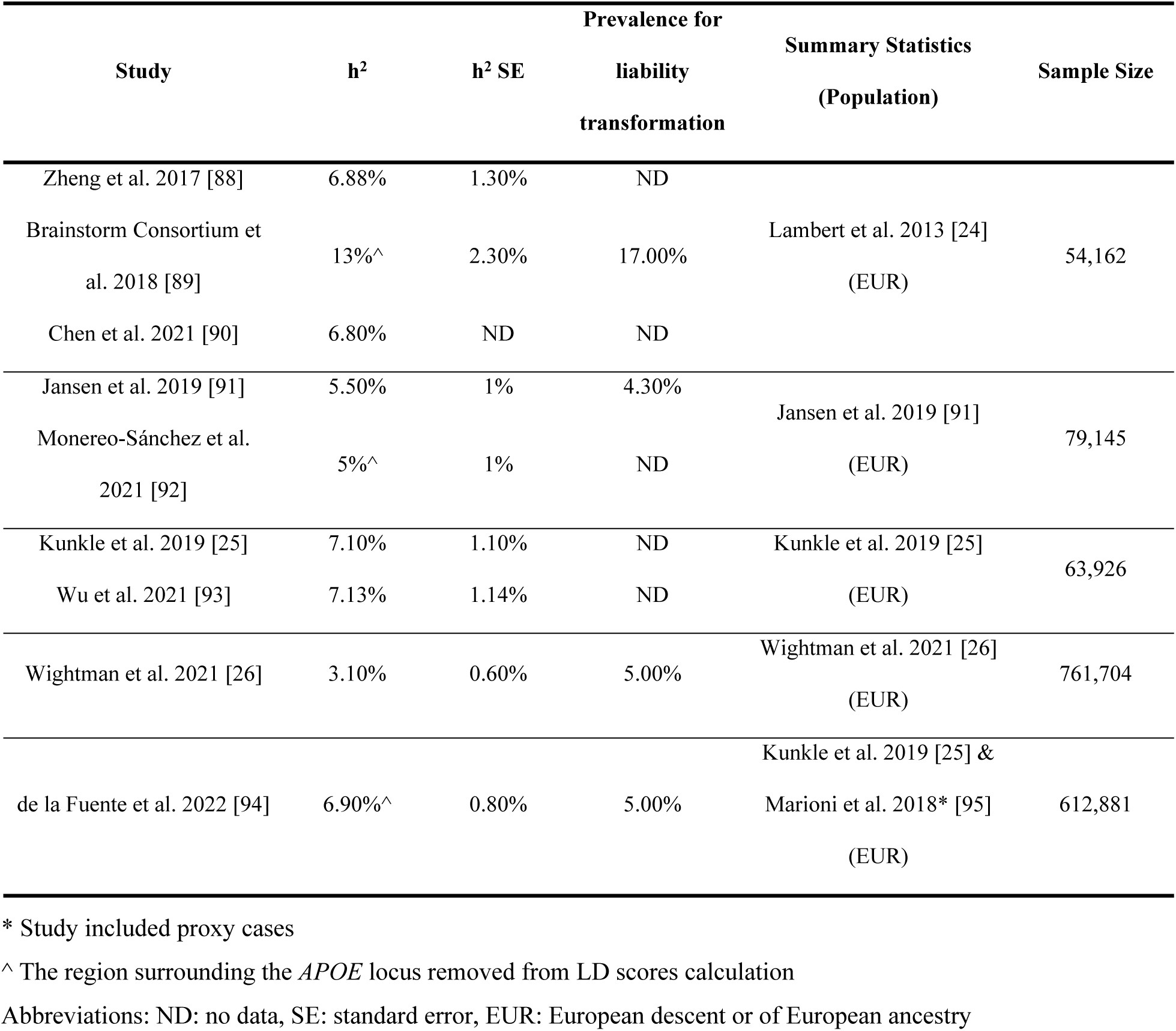
SNP-based LOAD heritability studies using LDSC.

Four major GWAS of LOAD have been pivotal in heritability estimation using LDSC (Table 3; Fig 3), all involving direct phenotyping of cases [24–26,91]. As discussed earlier, multi-consortia collaborative efforts revolutionized large-scale genetic studies of LOAD but also introduced heterogeneity into both study design and included populations, leading to variability in heritability estimates. Beginning with the LOAD GWAS published by Lambert et al. (2013), its stage 1 summary statistics formed the basis for three subsequent LOAD heritability estimates [88–90]. This foundational study involved a meta-analysis of four consortia (the ADGC, the Cohorts for Heart and Aging Research in Genomic Epidemiology (CHARGE) Consortium, the European Alzheimer’s Disease Initiative (EADI), and the Genetic and Environmental Risk in Alzheimer’s Disease (GERAD) Consortium) under the International Genomics of Alzheimer’s Project (IGAP), with a total of 54,162 participants in the analysis. Building on these efforts, Kunkle et al. (2019) leveraged the expanded IGAP study to become one of the largest GWAS of LOAD to date [25]. This comprehensive study incorporated 46 cohorts from the IGAP, including 17 new ones, increasing the sample to 63,926 individuals. Two LOAD heritability estimates were based solely on the stage 1 discovery sample [25,93]. Jansen et al. (2019) performed GWAS utilizing not only IGAP data but also incorporating two additional independent consortia: the Psychiatric Genomics Consortium (PGC-ALZ) and the Alzheimer’s Disease Sequencing Project (ADSP), boosting the total sample size to 79,145 [91,92]. Wightman et al. (2021) further expanded the scope of LOAD genetic research by including additional cohorts from Europe and the US not previously considered in Jansen et al. (2019), resulting in a sample size of 761,702 (43,725 cases, 717,979 controls) and its own heritability estimate based on this LOAD GWAS [26]. All of these large GWAS included cases and controls directly phenotyped for LOAD status.

In contrast to the direct LOAD phenotyping approaches described above, Marioni et al. (2018) contributed a study using proxy-phenotypes based on family history (GWAX), leveraging data from large electronic health records available for UK Biobank participants ages 40 to 69 [95]. Using an innovative approach, de la Fuente et al. (2022) broadened the heritability estimation of LOAD by integrating both GWAS and GWAX while addressing the attenuated heritability estimates when directly combining the summary statistics from the two [94].

Beyond the heterogeneity across cohorts, variations in heritability estimates are observed even within studies using the same LOAD summary statistics. This variability could stem from the unique challenges in estimating heritability for binary traits like LOAD, as compared to quantitative traits. The process of estimating heritability for binary traits involves an initial estimation made on the observed scale, using the sample prevalence as the threshold followed by a transformation to the liability scale utilizing the population prevalence [96,97]. This approach results in a mix of observed- and liability-scale heritability estimates across studies, even when leveraging the same summary statistics, contributing significantly to the diverse heritability values reported in the literature. The impact of this methodological variation on the heritability estimates can be seen in studies leveraging summary statistics [24]. Zheng et al. (2017) and Chen et al. (2021) reported similar observed-scale heritability estimates of 6.88% and 6.80%, respectively [88,90]. In contrast, the Brainstorm Consortium et al. (2018) provided a markedly different liability-scale heritability estimate of 13%, using a population prevalence of 17% for the transformation [89]. Liability scale heritability is critical in adjusting for the ascertainment of the binary traits and making the heritability estimates comparable across studies using appropriate population prevalence for transformation. The challenge of interpreting and comparing these mixed-scale estimates is further compounded by the wide range of population prevalence figures used in liability-scale transformations. Across various studies, these prevalence estimates range from 4.3% to 17% [26,89,91,94]. This variability in prevalence estimates can significantly impact heritability figures, making direct comparisons between studies difficult.

### SNP-Based Heritability Re-Evaluation

In addition to the 16 SNP-based studies (Tables 2 and 3) included in this review that met our criteria (Fig 1), we identified several “re-evaluations” of LOAD SNP-based heritability where the heritability estimates were primarily generated in the context of gene discoveries. With variation in the data included, the methodology considered, and assumptions made, these re-evaluations based on previous studies resulted in a range of incomparable estimates. At least three SNP-based heritability re-evaluations studies have been published: one utilizing both GCTA and LDSC [98], one using only LDSC [99], and a third one employing LDAK [100], a method that accounts for minor allele frequency and LD when estimating a SNP’s influence on a trait’s heritability [101,102]. The number of datasets or studies included ranged from four [98] to ten [99] that resulted in a large range of heritability estimates with a consistent population prevalence of 5%: the LOAD heritability estimates using LDSC ranged from 9% to 17% [98] and 3% to 42% [99]. As expected, the GCTA estimates were higher at 25% to 31% [98]. LDAK estimates generally aligned more closely with GCTA, and estimates derived from LDAK were generally higher than LDSC. As an example, heritability estimates using data from Kunkle et al (2019) increased from 7% with LDSC (Table 3) to 21% using LDAK [100].

## Conclusion and Discussion

In this review we evaluated multiple measures of heritability for LOAD and showed that direct comparisons between heritability studies are difficult due to study population variability, methodological differences, and other factors outlined in this review. Understanding and critically evaluating heritability estimates require careful consideration of underlying assumptions, methodological nuances, and the challenges inherent in quantifying genetic contributions to complex diseases like LOAD. Another significant limitation of current LOAD heritability research is its predominant focus on European descent populations. This lack of diversity, well-documented in genetic studies [103], substantially impacts the generalizability and broader applicability of obtained estimates. However, it is of note that all of the studies we reviewed were of European ancestry and variation in heritability estimates was still huge. Moving forward, the recognition that there is no one real measure of heritability should be promoted, reinforcing that each estimate captures nested components of genetic architecture within the specific methodological and population context from which they were obtained. Clearly, future studies would benefit from increased population diversity, standardized phenotyping approaches, and careful consideration of both genetic and environmental factors to provide a more comprehensive understanding of LOAD heritability across diverse populations. Despite the limitations in estimating LOAD heritability, doing so is still important as LOAD is a neurodegenerative disorder with significant public health implications, and knowledge of heritability is a gateway to better understanding its genetic architecture and guiding future research efforts [3,67,68,104]. In this systematic review, we identified 23 studies providing LOAD heritability estimates, employing either twin-based (n=7) or SNP-based (n=16) approaches. The included studies demonstrated a range of heritability estimates that varied considerably (Fig 4), reflecting the diversity in study designs, estimation methodologies, and the underlying heterogeneity of LOAD. Family-based approaches that leverage the genetic similarities between relatives to estimate heritability, generally yielded higher estimates, ranging from 37% to 89% based on twin studies. The higher estimates for twin studies are expected as these implicitly include all genetic variants and their possible non-additive effects, while also potentially including shared environmental effects that are difficult to dissect. This potentially leads to an overestimation of heritability. In contrast, SNP-based approaches that compute heritability estimates based on measured genetic variants across the genome, provided relatively lower estimates (3.1% to 53%), as they are limited by what has been genotyped or imputed [59]. Among them, studies using summary statistics provided relatively lower estimates (LDSC-based; 3.1% to 13%) compared with those using genotyped or imputed data (GCTA-based; 19% to 53%). As we recognize that LDSC-based heritability estimates tend to be downwardly biased [87,102,105,106], in contrast to twin studies that can capture gene-environment interactions, along with other factors, that potentially inflate heritability estimates [107–109]. While acknowledging that the heritability of LOAD is variable to some extent [80], we posit that the heritability of LOAD in most European-descent populations falls between 40-60%, capturing the upper range of SNP-based heritability estimates and the lower range of twin-based estimates. We think that this range better represents the substance of LOAD heritability than any single point estimate from any given study.

**Fig 4.**
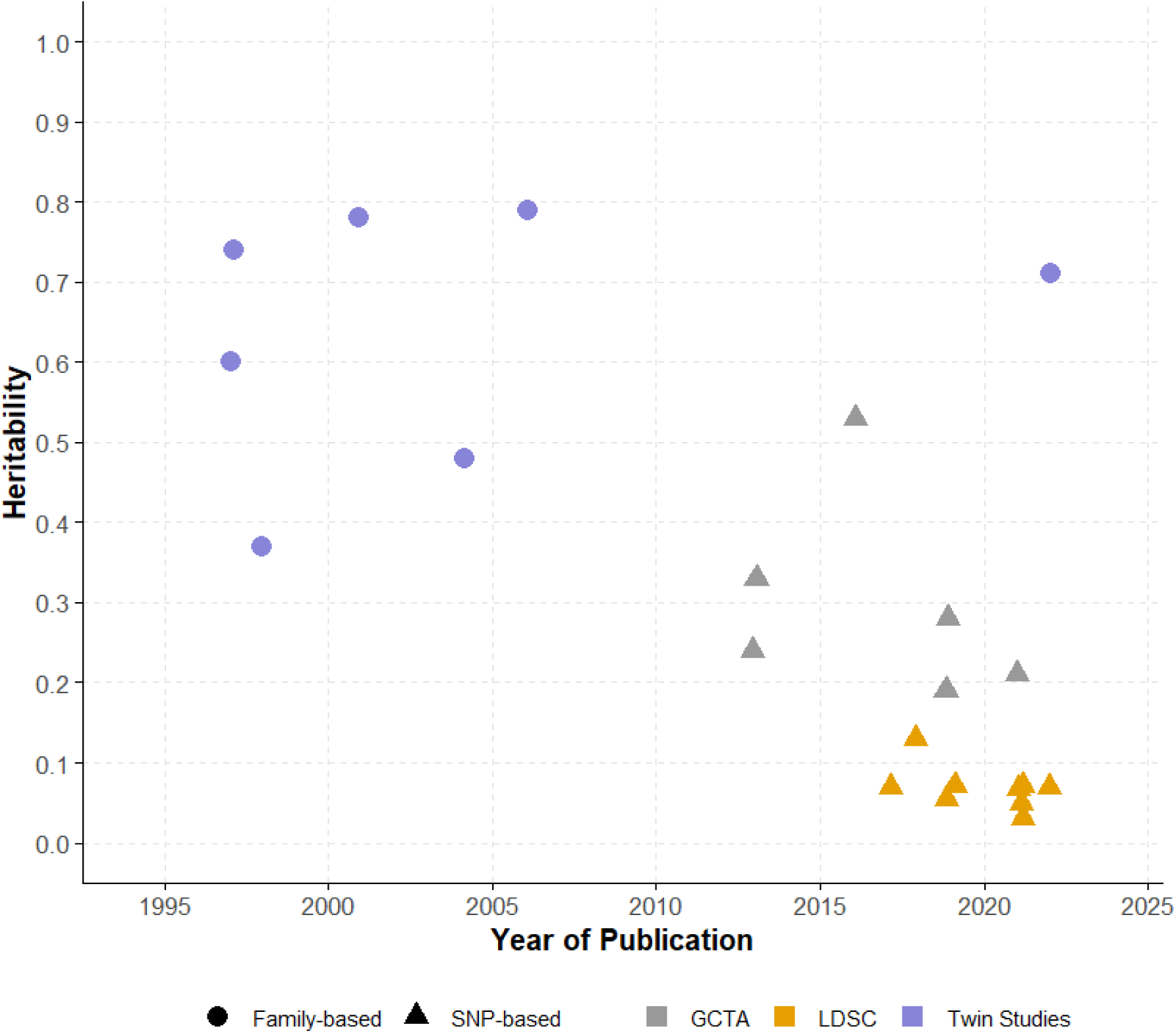
Heritability estimates across studies over time. Here shows the heritability estimates for LOAD from all studies identified, published between 1997 and 2022. Within the figure, each data point represents an individual estimate corresponding to the value on the y-axis, with shapes distinguishing between family-based (circle) and SNP-based (triangle) approaches, while colors indicating specific methods: purple for twin studies, gray for GCTA, and orange for LDSC.

## Data Availability

This is a systematic review, and all data is available from the references.

